# Differential associations between SARS-CoV-2 infection, perceived burden of the pandemic and mental health in the German population-based cohort for digital health research

**DOI:** 10.1101/2024.02.14.24302768

**Authors:** Lavinia A. Steinmann, Luise V. Claaß, Moritz Rau, Janka Massag, Sophie Diexer, Bianca Klee, Cornelia Gottschick, Mascha Binder, Daniel Sedding, Thomas Frese, Matthias Girndt, Jessica Hoell, Irene Moor, Jonas Rosendahl, Michael Gekle, Rafael Mikolajczyk, Nils Opel

## Abstract

**Introduction:** Understanding the potential adverse effects of the Covid-19-pandemic remains a challenge for public mental health. In this regard, the differentiation between potential consequences of actual infection with SARS-CoV-2 and the subjective burden of the pandemic due to measures and restrictions to daily life remains elusive.

**Methods:** Here we investigated the differential association between infection with SARS-Cov-2 and subjective burden of the pandemic in a study cohort of 7601 participants from the German population-based cohort for digital health research (DigiHero), who were recruited between March 4^th^ and April 25^th^ 2022. Data was collected using the online survey tool LimeSurvey® between March and October 2022 in consecutive surveys, which included questionnaires on infection status and symptoms following COVID-19 as well as retrospective assessment of the subjective burden of the pandemic.

**Results:** We observed an association of a past SARS-CoV-2 infection on deteriorated mental health related symptoms, whereas no association or interaction with burden of the pandemic occurred. The association was driven by participants with persistent symptoms 12 weeks after acute infection. On a symptom specific level, neuropsychiatric symptoms such as exhaustion and fatigue, concentration deficits as well as problems with memory function were the primary drivers of the association.

**Conclusion:** Our findings underscore the impact of SARS-CoV-2 infections on mental health in patients suffering from ongoing symptoms 12 weeks after infection. As the association between SARS-CoV-2 infection and mental health appeared more pronounced in populations with higher vulnerability for mental disorders, increased attention should be dedicated towards these subgroups regarding the prevention of infection.

## 1. Introduction

The aftermath of the COVID-19 pandemic continues to have a profound and multifaceted impact on public health and remains a major challenge for both the public and healthcare professionals. Particularly, a pronounced impairment in mental health has been discussed as a potentially long-lasting consequence of the COVID-19 pandemic. This conceivably arises from a combination of increased stress and anxiety, changes in routine, socioeconomic challenges, consequences of containment measures such as social isolation, ongoing neuropsychiatric symptoms after SARS-CoV-2 infection and an increase in the incidence of mental health disorders (Penninx et al., 2022).

A growing body of literature has therefore aimed to investigate the exact and differentiated impact of the COVID-19 pandemic on mental health. Soon after the pandemics begin in Germany in March 2020 an increase of self-reported psychosocial distress was observed in population-based studies compared to pre-pandemic levels (Giel et al, 2022). Meta-analyses pooling data from international studies reported anxiety and depression levels having the strongest increase during the pandemic (Vindegaar & Benros, 2022; Prati & Mancini, 2021; Robinson et al., 2022). Confirmed by data from the US Centers for Disease Control and Prevention (CDC) are direct correlations between the level of symptoms related to anxiety and depression and the average number of reported daily COVID-19 cases (Jia et al., 2021). The deterioration in mental health during the first wave of the pandemic was most likely due to disruptive societal changes, governmental restrictions and containment strategies rather than infection-associated with absolute COVID-19 cases still low (Robinson et al., 2022). Subsequently, amidst the sustained progression of the pandemic and the continuing rise in infection rates, an association between SARS-CoV-2 infections and an increased risk for new-onset of mental disorders was observed in a longitudinal study based on electronic health record (EHR) data of >89 million patients (Taquet et al., 2022). Similarly, recent publications from 2023 as well as pre-pandemic data from a nationwide registry study in Denmark substantiate an increased risk for the development of neuropsychiatric disorders following respiratory infection compared to the general population, albeit with similar rates for COVID-19 and non-COVID-19 related respiratory infections (Benros et al., 2013; Gronkjaer et al., 2023; Nersesjan et al., 2023).

A subset of patients with COVID-19 develop post-acute COVID Syndrome (PACS), which can include neuropsychiatric symptoms, such as fatigue, cognitive impairment, sleep disturbances, symptoms of post-traumatic stress disorder, concentration problems, and headache (Crook et al., 2021; Vindegaard & Benros, 2020). A large meta-analysis summarizing 51 studies with 18.917 patients and a mean follow-up of 77 days found sustained levels of neuropsychiatric symptoms in patients after recovery from the acute infection, foremost sleep disturbance, fatigue and cognitive impairment (Badenoch et al., 2022).

Concerning the impact of the COVID-19 pandemic on mental health, it is crucial to distinguish between at least two distinct factors contributing to the observed decline in mental well-being in the course of the pandemic. Firstly, there is growing evidence supporting the notion that infection with SARS-CoV-2 can lead to persistent neuropsychiatric sequelae in some patients, such as fatigue, depression, and cognitive impairment, through neurobiological pathways not yet fully elucidated, and is associated with an increased risk of new onset of mental illnesses. Secondly, the pandemic-related containment measures have brought about significant alterations in the daily lives of a substantial portion of the global population (Mehandru & Merad, 2022; Premraj et al., 2022). These psychosocial changes include heightened social isolation, changes in the work environment, and reduced engagement in recreational activities. These alterations might impose an additional burden on public mental health.

In order to adjust preventive measures and inform policy makers for future pandemics, it thus appears highly relevant to clarify the relationship between both factors and impairment in mental health. Despite this notion, the majority of previous studies did not differentiate between the potential impact of SARS-CoV-2 infection and the impact of the pandemic related burden on everyday life due to control measures and restrictions. Hence, the present study aimed to differentially investigate the potential association between mental health related symptoms and SARS-CoV-2 infection as well as the subjective burden of the COVID-19 pandemic. To this end, we used data from the DigiHero study, an ongoing nationwide, population-based digital study in Germany (Diexer et al., 2023; Massag et al., 2023). We hypothesized that both SARS-CoV-2 infection as well as the retrospectively rated subjective burden of the COVID-19 pandemic would be related to mental health related symptoms.

## 2. Methods

### 2.1. DigiHero Study and Data assessment

We study used data from the prospective cohort study for digital health research in Germany (DigiHero, DRKS Registration-ID: DRKS00025600) that has been described in detail elsewhere (Diexer et al., 2023; Massag et al., 2023). The cohort was initialized in January 2021 in Halle (Saale), Germany, and since then stepwise expanded to the states of Saxony-Anhalt, Saxony and Thuringia and several other regions nationwide. Persons aged 18 - 85 years were randomly drawn from resident registries and received a postal invitation. The registration for the study and the subsequent answering of questionnaires was conducted digitally. Immediately after registration, a baseline questionnaire asking for sociodemographic characteristics was sent to the participants via email using the online survey tool LimeSurvey®.

Data on somatic and mental health symptoms as well as subjective burden of the pandemic were collected between August 29^th^ and October 5^th^ 2022 at the end of the seventh phase of the pandemic with relatively low case numbers for acute infections and eased restrictions (Tolksdorf et al., 2022). During this time the perception was widely spread that the pandemic is overcome and ending, resulting in calling our assessment “post-pandemic assessment”. Sociodemographic baseline characteristics of the study population including age, sex, and education were obtained at the initial recruitment between March 4^th^ and April 25^th^ 2022. Data was analyzed using IBM SPSS Statistic (version 29).

### 2.2. Assessment of SARS-CoV-2 infection anamnesis

Participants were asked at the baseline assessment to indicate if they were ever tested positive for SARS-CoV-2. In addition, we collected data on number and type of test as well as symptoms of acute infection. Participants were defined as having been infected with the SARS-CoV-2 virus if they had had a positive PCR, a positive rapid test confirmed by a PCR test, multiple positive rapid tests or a positive rapid test and symptoms of acute infection.

Participants reporting a SARS-CoV-2 infection at the baseline recruitment were asked for symptoms 4-12 weeks and over 12 weeks after infection in a follow up survey. Between July 8^th^ and August 12^th^ 2022 the study cohort was asked about new SARS-CoV-2 infections in the meantime, followed by the questionnaire on persisting symptoms sent out on October 19^th^. In addition, time in weeks between SARS-CoV-2 infection and the post-pandemic assessment infection was assessed.

### 2.3. Assessment of Subjective Burden of the Pandemic and Life Satisfaction

To measure the subjective burden of the COVID-19 pandemic, the post-pandemic assessment covered a questionnaire asking participants to retrospectively rate the subjective burden of specific restrictions in daily life during the last 2 years of the pandemic. The questionnaire included a total of 24 questions on diverse sections, among others school and shop closures, work related restrictions, obligation to wear a face mask, travel restrictions, restrictions with regard to recreational activities **(see supplements S1 for detailed questionnaire)**. Participants were asked to rate the presence and severity of each symptom in retrospective throughout the pandemic, i.e. approx. the last two years, by using a 10-point likert-scale for each item with possible ratings of the subjective burden ranging from “1” to “10” with higher values representing higher subjective burden. Missing values were imputed using each item’s mean value with a frequency of missing values being less than 4% for all items. For further analyses, we calculated an overall sum score covering responses from all 24 items. The calculated overall subjective burden sum score showed excellent internal consistency (alpha= .933, n=24). In addition to the retrospective rating of subjective burden of the pandemic, participants were asked to rate the current level of satisfaction with their life using a single question with a likert-scale response ranging from “1” to “10” with higher values representing higher satisfaction.

### 2.4. Assessment of Mental Health related symptoms

The post-pandemic assessment included a questionnaire on somatic and mental health symptoms including 27 questions on diverse sections of health including symptoms related to mental health, acute infection as well as cardiovascular and gastrointestinal disorders, called “overall symptom load” **(see supplements S1 for detailed questionnaire)**. Participants were asked to rate the presence and severity of each symptom by using a 6-point likert-scale for each item with possible ratings of the symptom severity ranging from “Not at all” to “Very severe”. Participants were also provided with the option to indicate if they were not sure what to reply to a question. Missing values were imputed using each item’s mean value with a frequency of missing values being less than 3% for all items. For further analyses, we calculated an overall sum score covering responses from all 27 items, as well as a mental health subscale covering responses from the seven mental health related items, namely symptoms of depression, anxiety, concentration problems, memory problems, sleep disorder, fatigue, and exhaustion (“mental health score”). Both the calculated overall symptom sum score (alpha= .894, n= 27) as well as the mental health subscale (alpha= .869, n=7) showed good to excellent internal consistency. A factor analysis using principal component analysis with oblique rotation confirmed the outlined approach by indicating a factor structure with all mental health symptoms loading primarily on the first factor explaining 27.7% of variance in all reported symptoms.

### 2.5. Statistical analyses

After identification of participants with and without SARS-CoV-2 infection, we performed a non-parametric Mann Whitney U Test regarding current satisfaction with life. Additionally, we performed regression analyses with current satisfaction with life as independent variable and overall symptom load, mental health score and subjective burden of pandemic.

We fitted a general linear model including SARS-CoV-2 infection status (having had a SARS-CoV-2 infection yes/no) and subjective burden of pandemic sum score as predictors and mental health score as dependent variable, adjusting for age, sex, and education. We also performed an interaction analysis between infection status and subjective burden of pandemic.

For further analyses, we replaced the dichotomous variable “infection status” by a more detailed variable with four groups: No reported infection; Infection without persistent symptoms; Infection followed by symptoms in 4-12 weeks but not >12 weeks after infection; Infection followed by symptoms >12 weeks after infection. For this analysis, we only included participants with infection whose infection dated back 12 weeks and more so that symptoms in this time window could have been assessed. In our analysis, we firstly looked at main effect of these groups, followed by post-hoc analyses of differences in mean mental health symptom load between the four groups. Subsequently, we investigated the effect size computing partial eta square for each mental health symptom.

## 3. Results

### 3.1. Characteristics of the study sample

The present analyses included a study population of n=7601 participants (mean age: 51.07, SD=15.00, age range 18-88; 61.2% female), **see Table 1 for detailed overview of participant characteristics**.

**Table 1.**
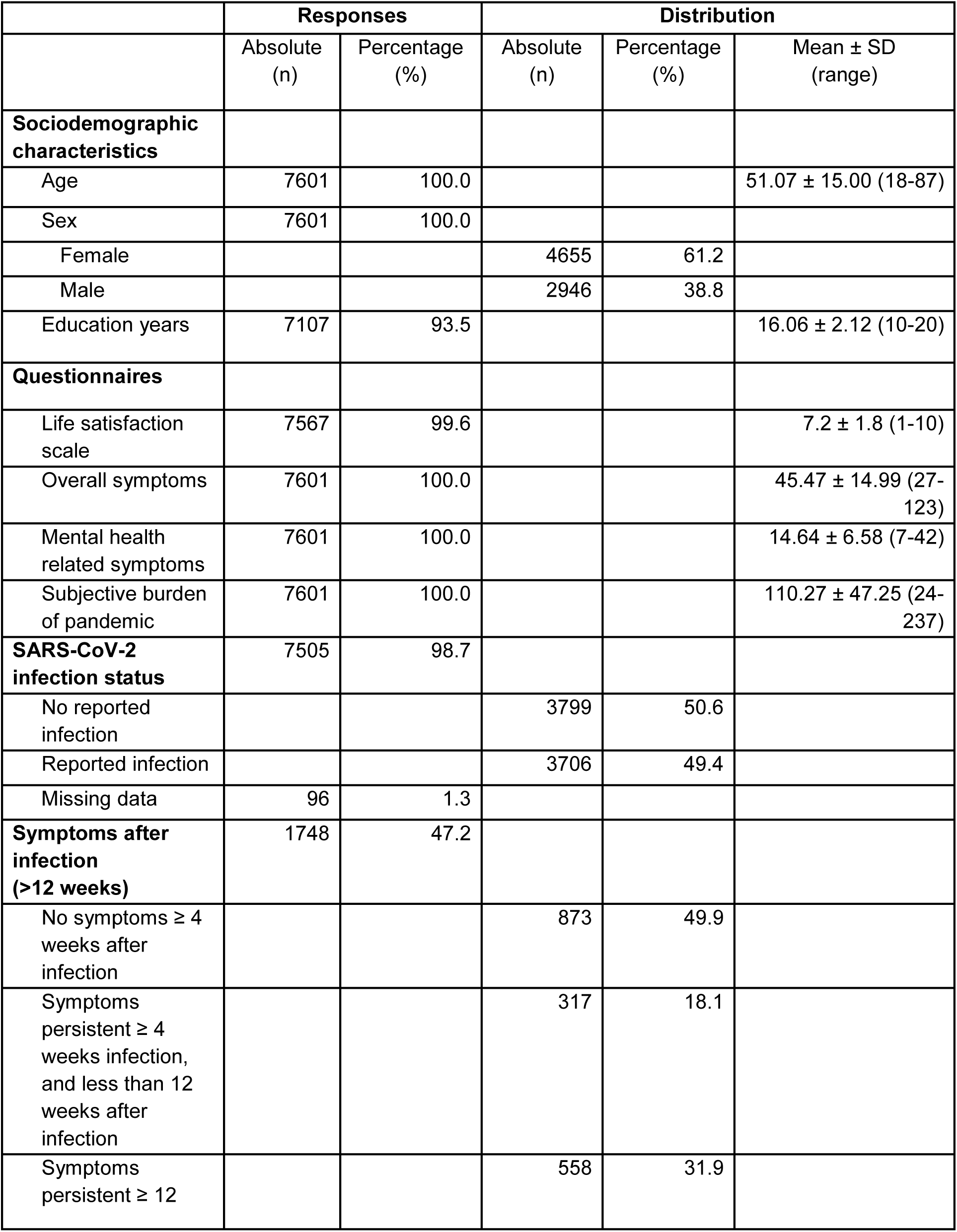
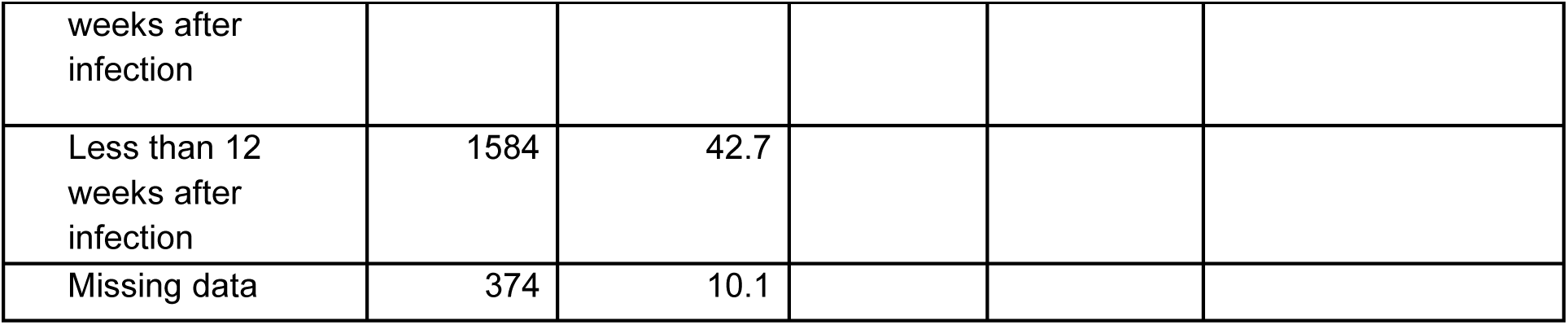
Sociodemographic characteristics, questionnaires and SARS-CoV-2 infections status of our study sample consisting of n=7601 participants.

48.8% (n= 3706) of the study population reported a previous infection with SARS-CoV-2. Participants with and without a history of SARS-CoV-2 infection did not report differences in current satisfaction with life as assessed using non-parametric Mann Whitney U Test (p= .864, Z= .172). Current satisfaction with life was associated with mental health symptoms (r= -.463, p <.001) and overall health symptoms (r= - .416, p <.001), but only weakly associated with subjective burden of the pandemic (r= -.088, p <.001) as assessed using Spearman Rank correlation analyses.

In 1748 of 3706 (47.2%) persons with reported SARS-CoV-2-infections, the infection dated back 12 weeks and more. Out of these study participants 18.1% (n= 317) reported COVID-19 related symptoms 4 to 12 weeks but not >12 weeks following infection and 31.9% (n= 558) reported COVID-19 related symptoms 12 weeks or longer following infection whereas 49.9% (n= 558) did not suffer from persisting symptoms.

### 3.2. Association between SARS-CoV-2 Infection, subjective burden of the pandemic and mental health

The general linear model including both infection status (having had a SARS-CoV-2 infection +/-) and subjective burden of pandemic as predictors and the mental health score as dependent variable indicated a small effect of previous SARS-CoV-2 infection on reported mental health symptoms (F(1, 7505)= 5.304, p= .021, ηp^2^= .001; M(infection+)= 14.84, SD (6.62), M(infection-)= 14.44, SD (6.54)). No significant association between subjective burden of the pandemic and mental health symptoms could be observed (F(1, 7505)= .944, p= .861). There was also no interaction effect of SARS-CoV-2 infection x burden of the pandemic on mental health symptoms (F(1, 7505)= 1.009, p= .451).

The model furthermore indicated significant main effects of sex (F(1, 7505)= 165.903, p< .001, ηp^2^= .025) and education (F(1, 7019)= 10.956, p< .001, ηp^2^= .002) on mental health symptoms, whereas no significant main effect of age was observed (F(1, 7505)= .050, p= .824).

We observed a significant interaction effect with a small effect size between SARS-CoV-2 infection and age (F(1, 7505)= 14.933, p< .001, ηp^2^= .002) on mental health symptoms **(see supplemental figure for detailed results)**. No significant interaction of SARS-CoV-2 infection and sex (F(1, 7505)= .272, p= .602) or SARS-CoV-2 infection and education could be observed (F(1, 7019)= 1.462, p= .227).

In the subsample consisting of four groups (No reported infection; Infection without persistent symptoms; Infection followed by symptoms in 4-12 weeks but not >12 weeks after infection; Infection followed by symptoms >12 weeks after infection), we could also detect a main effect of infection on mental health symptoms (F(1, 5547)= 69.3, p< .001, ηp^2^= .046). Post-hoc analysis indicated that individuals who reported symptoms more than 12 weeks after infection showed the most pronounced symptom load, while the other three studied groups had similar levels of mental health symptoms **(see Figure 1)**. Also in this analysis, subjective burden of the pandemic demonstrated neither association nor interaction with the mental health sum score.

**Figure 1.**
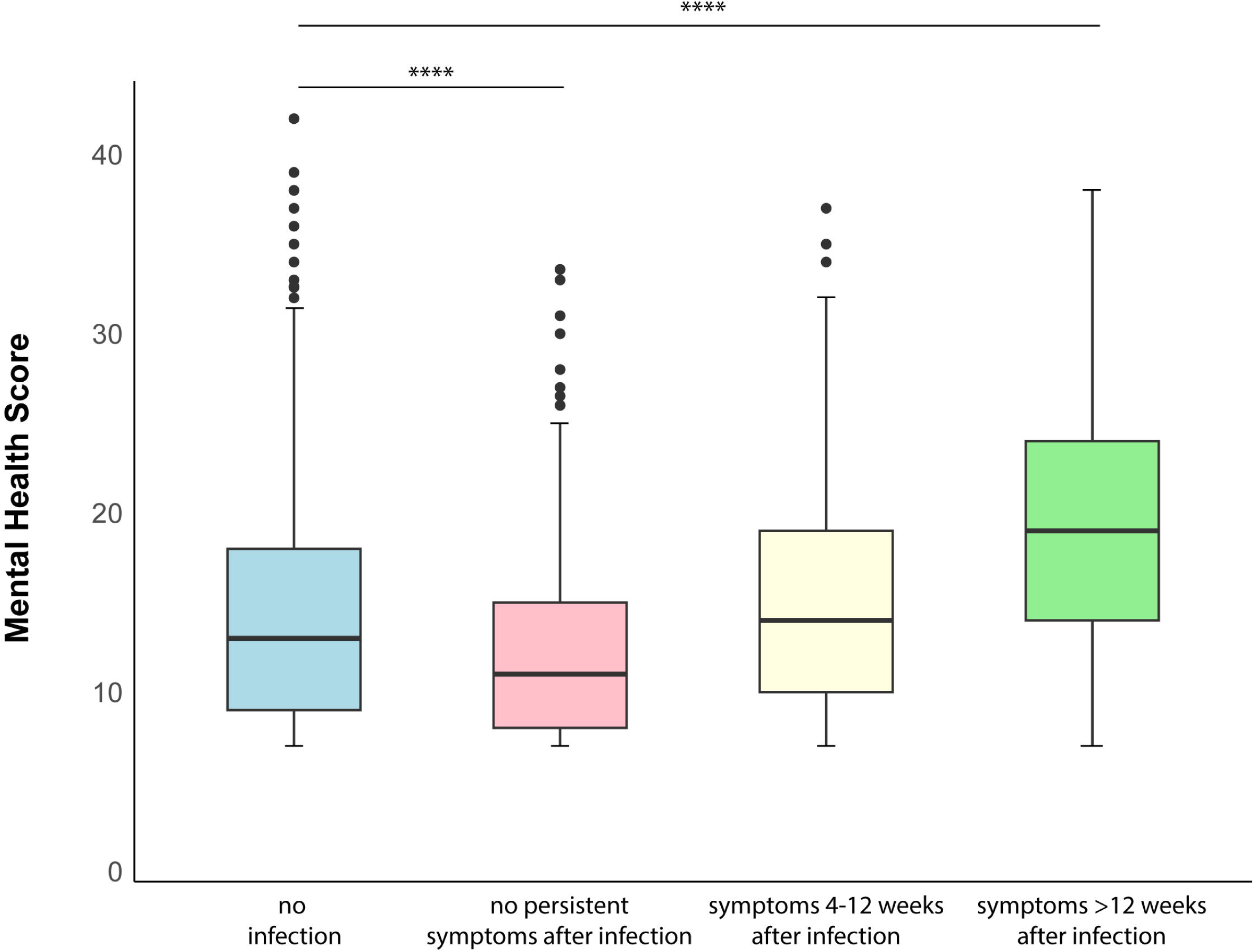
Boxplots depicting distribution of mental health related symptoms according to infection status. Mental health score ranging from 7-42 with higher values indicating increased symptom load. As a statistical test a non-parametric Kruskal-Wallis test using a Bonferroni correction with a confidence interval of 95% was used. (****) p ≤ 0.001.

On a symptom specific level, neuropsychiatric impairment involving exhaustion, fatigue, concentration problems and problems with memory function were the primary drivers of association between SARS-CoV-2 infection and mental health symptoms showing the largest effect sizes **(see Figure 2)**.

**Figure 2.**
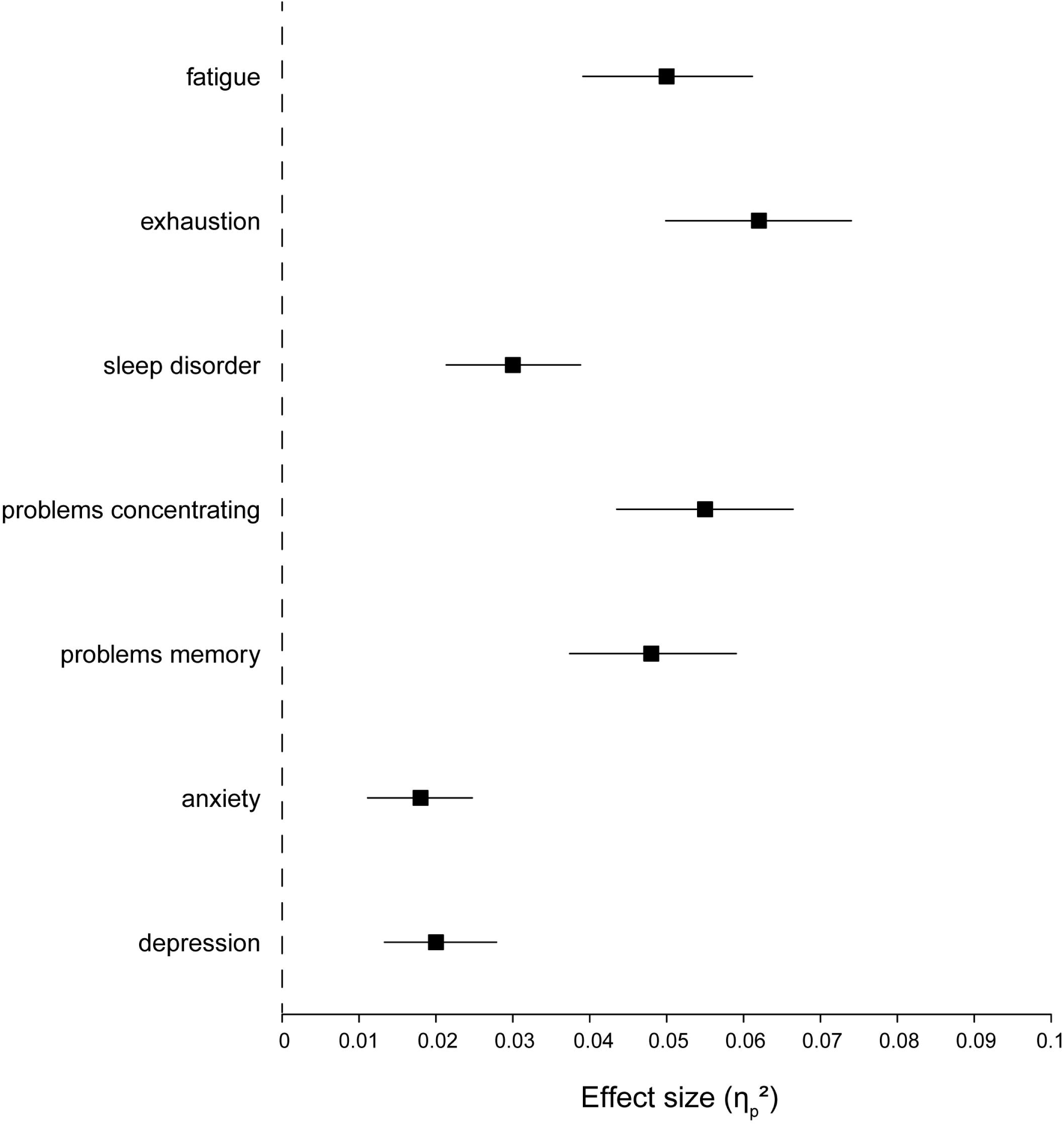
Effect sizes of mental health related symptoms. Partial eta squared (ηp^2^) values are shown as effect sizes of the different mental health related symptoms as possible drivers for the association of a SARS-CoV-2 infection and mental health with a confidence interval of 95%. Values ≥ to 0.01 indicate a small effect, values ≥ 0.06 indicate a medium effect and values ≥ 0.14 indicate a large effect.

## 4. Discussion

In the present study, we identified an association between SARS-CoV-2 infection and mental health symptoms, distinguishing it from the subjective burden of the pandemic-related measures. It is noteworthy that even after considering the impact of perceived pandemic burden, the link between infection and mental health persisted. Notably, this association was driven by a subgroup characterized by persisting symptoms over 12 weeks after infection, and it was primarily manifested through neuropsychiatric symptoms such as exhaustion, concentration difficulties, and memory impairment. Well-known demographic vulnerability factors such as lower education and female gender were associated with deteriorated mental health. These findings underscore the relationship between SARS-CoV-2 infection and potential long-lasting symptoms such as mental health deterioration and hold several relevant implications for future research.

Our finding of an association between SARS-CoV-2 infection and mental health impairment is well in line with a growing body of literature corroborating the influence of respiratory infections on neuropsychiatric impairment. Regarding SARS-CoV-2 infection specifically, recent large-scale evidence from nationwide register studies from Denmark demonstrated an elevated risk for the development of psychiatric and neurological disorders following infection (Grønkjær et al., 2023; Nersesjan et al., 2023). These findings are supported by large-scale EHR based studies confirming an increased risk for neurological and psychiatric disorders following SARS-CoV-2 infection (Taquet et al., 2022).

Our findings build upon the aforementioned evidence and extend it by incorporating an assessment of the subjective burden associated with pandemic-related measures. Importantly, our results affirm that the connection between SARS-CoV-2 infection and mental health impairment is independent of this subjective burden.

These findings are further supported by pre-pandemic studies indicating an association between infectious disorders including respiratory infections and elevated risk for the development of psychiatric disorders (Benros et al., 2013). Differentiated analyses of pandemic data indicated that the association between respiratory infections and mental health was similarly present for SARS-CoV-2 and non-SARS-CoV-2 related infections (Nersesjan et al., 2023). Likewise, the pre-pandemic and pandemic studies consistently demonstrated that the risk for mental health impairment following infection increases with infection severity (Köhler et al., 2017; Nersesjan et al., 2023).

Taken together, the existing evidence suggests an association between infections and mental health impairment that is not specific for SARS-CoV-2 and is moderated by infection severity. The latter notion might also explain the small effect size that we observed for the association between SARS-CoV-2 infection and mental health in the present study. As our study built on data from the general population and did not take into account infection severity, it appears plausible to assume that the present sample included a large number of participants with mild infections.

The symptoms that were primarily driving the association between infection and mental health impairment were exhaustion, fatigue, concentration and memory deficits. Again, this finding is well in line with findings from studies reporting concentration deficits, amnesia and fatigue among the most prevalent symptoms in the long-term course of following infection (Ceban et al., 2021; Kim et al., 2023). Indeed, cognitive dysfunction, fatigue and memory loss are now widely considered as defining core symptoms of post-infectious neuropsychiatric sequlae following infection with SARS-Cov-2 also referred to as Long and Post COVID syndrome (Davis et al., 2023).

Low levels of education as a well-known vulnerability factor for psychiatric disorders was associated with worse mental health. Furthermore, we could confirm a main effect of sex with women being more impaired than men concerning mental health. Additionally, the cohort-based approach of the present study allowed us to analyze potential moderating factors. Results from these analyses indicated that risk for mental health impairment following infection differed between subpopulations depending on age, potentially being a relevant factor for more severe infections per se.

This underscores the significance of socioeconomic factors as potential risk indicators for post-infection mental health outcomes, warranting heightened attention to this particular subpopulation. While this aligns with established evidence indicating increased vulnerability to mental disorders in low socioeconomic status populations, the mechanisms through which infections exert a more pronounced impact in this demographic remain unclear. One plausible explanation is that lower general health within this group may contribute to this observation, though this remains speculative as our study lacked specific data on general health status before SARS-CoV-2 infection. To address this gap, future research should elucidate the potential roles of both psychosocial and biological factors, exploring risk and resilience dynamics, as well as somatic comorbidities that could modulate the relationship between infection and mental health. This comprehensive approach is essential for a more nuanced understanding of the interplay between socioeconomic status, infection, and mental health, facilitating targeted interventions and support for at-risk populations.

The moderating role of age should also be noted with increasing age conferring a higher risk for mental health impairment following infection. Thus, our findings affirm the importance of aging processes in the context of mental health impairment following infection and suggest higher age as a risk factor. Again, as we did not have further information on aging related factors, the underlying mechanisms remain elusive.

Our study comes with strengths and limitations. Strengths include the combination of a relatively large sample size with information on infection status and subjective burden of pandemic related measures thus allowing us to disentangle between the infection and burden of the pandemic-related measures. The cohort-based methodology, relying on self-reports, stands out as another strength, enabling a nuanced exploration of the association between infection and mental health at a symptom-specific level. This approach complements existing large-scale studies that rely on registers or HER-data (Benros et al., 2013; Taquet et al., 2022). Limitations comprise the limited depth of information on infection severity as a potential moderator of the observed association between infection and mental health. Furthermore, as the study was based on self-reports in the general population, we did not have the possibility to validate the observed symptoms based on external ratings or health records. The rating of the subjective burden of the pandemic comprised the last 2 years during the pandemic, potentially provoking a recall bias as a common problem in studies relying on self-reports. Finally yet importantly, the questionnaire on persisting symptoms after infection was not sent out individually after a distinct time after confirmed SARS-CoV-2 infection lacking standardization.

## 5. Conclusion

In summary, our study affirms a link between SARS-CoV-2 infection and mental health impairment, independent of the subjective burden associated with pandemic-related measures. A subgroup with persistent symptoms 12 weeks after infection displays significant deterioration of mental health. Interestingly, the main impairments are related to neuropsychiatric symptoms such as exhaustion, memory impairment and concentration deficits. Moreover, our investigation identifies moderating factors, indicating that individuals with low socioeconomic status and higher age may be at a heightened risk for developing mental health impairment post-infection. These findings emphasize the need for future research to elucidate the biological mechanisms that underlie this relationship, paving the way for the development of innovative preventive measures and therapeutic interventions. This insight holds promise for advancing our understanding and ultimately addressing the mental health implications of SARS-CoV-2 infection.

## Data Availability

All data produced in the present study are available upon reasonable request to the authors.

## Acknowledgements

There was no specific funding for this analysis. The DigiHero study is funded by internal resources of the Medical Faculty of the Martin Luther University Halle-Wittenberg and part of the recruitment was co-funded by the Ministry of Economy, Science and Digitalization of the Federal State of Saxony-Anhalt (Germany).

## Ethical standards

The Ethics Committee of the Martin Luther University Halle-Wittenberg (Registration number 2020-076) approved the study. Informed consent was obtained from all participants. The authors assert that all procedures contributing to this work comply with the ethical standards of the relevant national and institutional committees on human experimentation and with the Helsinki Declaration of 1975, as revised in 2008.

## Appendices

**A.1 Questionnaire “postpandemic assessment”** covering assessment of somatic and mental health symptoms.

**A.2 Questionnaire “postpandemic assessment”** covering the subject burden of the pandemic.

**Figure A.1** displaying interaction between SARS-CoV-2 infection status and age.

