## Appendices for "Differential associations between SARS-CoV-2 infection, perceived burden of the pandemic and mental health in the German population-based cohort for digital health research"

**A.1** Questionnaire on somatic and mental health symptoms (translated from German)

**Do you suffer currently, i.e. within the last four weeks, from the hereafter-mentioned symptoms and if so, how was the severity of these symptoms?**

Please choose the answer that applies to you for each item:

|  | **Not**  **at all** | **Very**  **weak** | **Weak** | **Medium** | **Severe** | **Very**  **severe** | **Not**  **sure** |
| --- | --- | --- | --- | --- | --- | --- | --- |
| **Fever** |  |  |  |  |  |  |  |
| **Lymph node swelling** |  |  |  |  |  |  |  |
| **Smell disorder** |  |  |  |  |  |  |  |
| **Taste disorder** |  |  |  |  |  |  |  |
| **Joint, muscle or limb pain** |  |  |  |  |  |  |  |
| **Fatigue** |  |  |  |  |  |  |  |
| **Exhaustion** |  |  |  |  |  |  |  |
| **Sleep disorder** |  |  |  |  |  |  |  |
| **Night sweats** |  |  |  |  |  |  |  |
| **Problems concentrating** |  |  |  |  |  |  |  |
| **Memory problems** |  |  |  |  |  |  |  |
| **Anxiety** |  |  |  |  |  |  |  |
| **Depression** |  |  |  |  |  |  |  |
| **Headache** |  |  |  |  |  |  |  |
| **Cold** |  |  |  |  |  |  |  |
| **Conjunctivitis** |  |  |  |  |  |  |  |
| **Earache or tinnitus** |  |  |  |  |  |  |  |
| **Shortness of breath** |  |  |  |  |  |  |  |
| **Sore throat** |  |  |  |  |  |  |  |
| **Cough** |  |  |  |  |  |  |  |
| **Tightness in the chest / cheastpain** |  |  |  |  |  |  |  |
| **Heart trouble like heart palpitations or cardiac arrhythmia** |  |  |  |  |  |  |  |
| **Vertigo** |  |  |  |  |  |  |  |
| **Stomach ache** |  |  |  |  |  |  |  |
| **Stomach flu/ diarrhoea** |  |  |  |  |  |  |  |
| **Nausea** |  |  |  |  |  |  |  |
| **Premenstrual syndrome (PMS) / period pains** |  |  |  |  |  |  |  |

**A.2** Questionnaire on burden of the pandemic (translated from German)

**How do you retrospectively rate the restrictions and burdens during the pandemic? Please indicate how much you felt restricted/burdened by the following measures taken during the Covid-19 pandemic! If certain items do not apply, please indicate, that did not feel restricted/burdened at all.**

Please choose the answer that applies to you for each item

1 – not restricted / not burdened at all

10 – very much restricted / very much burdened

|  | **1** | **2** | **3** | **4** | **5** | **6** | **7** | **8** | **9** | **10** |
| --- | --- | --- | --- | --- | --- | --- | --- | --- | --- | --- |
| **Preschool children could not attend nursery** |  |  |  |  |  |  |  |  |  |  |
| **School children could not attend school** |  |  |  |  |  |  |  |  |  |  |
| **Trainees/ students could not be taught in presence** |  |  |  |  |  |  |  |  |  |  |
| **Wearing face masks in shops** |  |  |  |  |  |  |  |  |  |  |
| **Wearing face masks in public transport** |  |  |  |  |  |  |  |  |  |  |
| **Social restrictions regarding friends** |  |  |  |  |  |  |  |  |  |  |
| **Social restrictions regarding family outside of household** |  |  |  |  |  |  |  |  |  |  |
| **No attendance of important family gatherings (e.g. wedding, funeral)** |  |  |  |  |  |  |  |  |  |  |
| **Social restrictions regarding colleagues** |  |  |  |  |  |  |  |  |  |  |
| **Closing of shops who do not sell convenience goods** |  |  |  |  |  |  |  |  |  |  |
| **Closed restaurants** |  |  |  |  |  |  |  |  |  |  |
| **Closed barbershops** |  |  |  |  |  |  |  |  |  |  |
| **No cultural events** |  |  |  |  |  |  |  |  |  |  |
| **No sport events** |  |  |  |  |  |  |  |  |  |  |
| **No possibility to do sports with others (e.g. in a sports club)** |  |  |  |  |  |  |  |  |  |  |
| **Limited radius of movement (up to 15 km)** |  |  |  |  |  |  |  |  |  |  |
| **Dusk-to-dawn curfew** |  |  |  |  |  |  |  |  |  |  |
| **Restrictions regarding foreign travel** |  |  |  |  |  |  |  |  |  |  |
| **Restrictions regarding domestic travel** |  |  |  |  |  |  |  |  |  |  |
| **Prohibition to visit patients in a hospital** |  |  |  |  |  |  |  |  |  |  |
| **Prohibition to visit residents in a nursing home** |  |  |  |  |  |  |  |  |  |  |
| **Necessity to work in home-office** |  |  |  |  |  |  |  |  |  |  |
| **Restrictions to go to physicians or other medical staff** |  |  |  |  |  |  |  |  |  |  |

### **Figure A.1** displaying interaction between SARS-CoV-2 infection status and age.

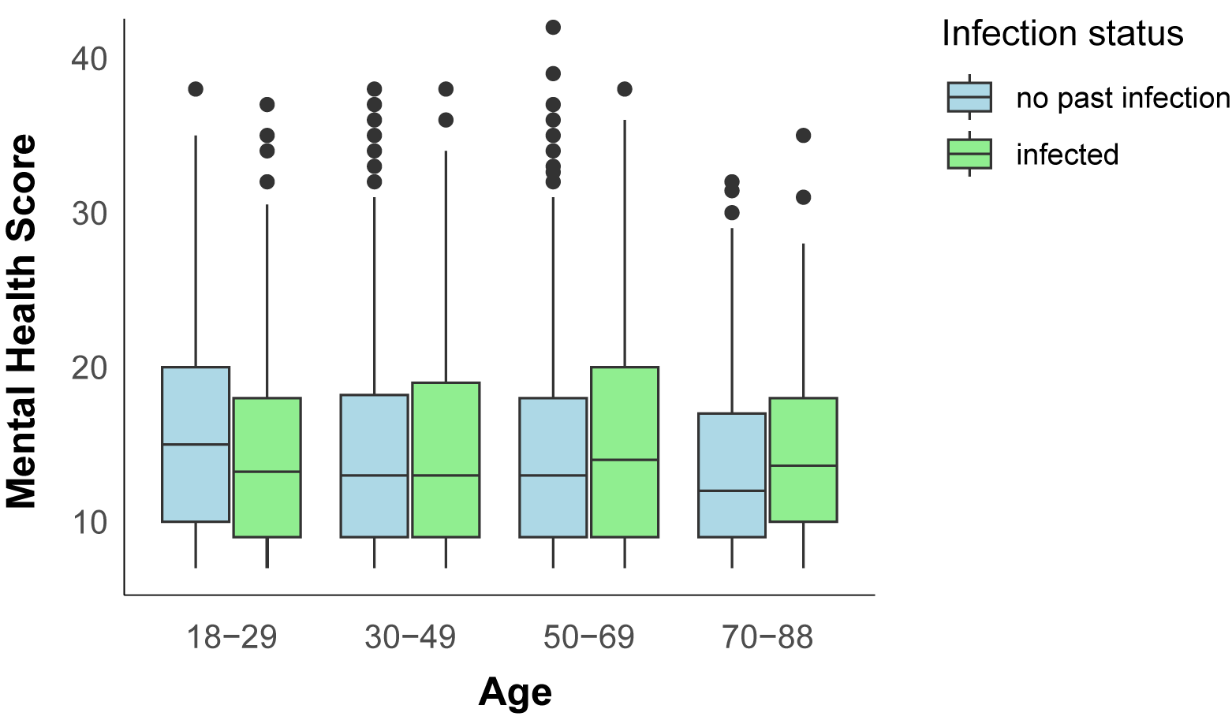
